# Promoting Resilience in Healthcare Workers during the COVID-19 Pandemic with a Brief Online Intervention

**DOI:** 10.1101/2021.07.28.21261304

**Authors:** NR DeTore, L Sylvia, ER Park, A Burke, JH Levison, A Shannon, KW Choi, FA Jain, DC Coman, J Herman, R Perlis, M Fava, DJ Holt

## Abstract

**Introduction:** The psychological wellbeing of healthcare workers has been impacted by the high levels of stress many have experienced during the COVID-19 pandemic. This study aimed to examine the feasibility and acceptability of a brief online course focused on introducing skills that could increase resilience and decreases emotional distress in healthcare workers during the pandemic.

**Materials and Methods:** Employees of a large healthcare system completed a survey at baseline, one month, and two months later. The online course, called Resilience Training for Healthcare Workers, consists of three 12-20 minute videos focused on evidence-based skills that support aspects of emotional resilience: mindfulness, mentalization, and self-compassion.

**Results:** A total of 554 participants completed the baseline survey, endorsing moderate to high levels of emotional distress. Of those who completed all three assessments and participated in the course (n = 38), significant improvements in resilience and reductions in emotional distress were found across two months, in comparison to those who did not participate in the course.

**Discussion:** These findings suggest that a brief, online intervention can improve the mental health of healthcare workers during a crisis such as the COVID-19 pandemic.

## Introduction

The World Health Organization has stated that supporting the mental wellbeing of healthcare workers during the COVID-19 pandemic is a critical priority (World Health Organization, 2020). During the pandemic, this population has experienced persistently elevated levels of stress related to the greatly increased clinical demands placed on them, during a time when they may also lack their typical sources of support. The psychological impact of the pandemic on this population was first observed in healthcare workers in Wuhan, China, where 45-50% reported depression and anxiety and 71% reported increases in emotional distress (Lai et al., 2020). Consistent with this finding, a systematic review of 44 studies conducted during the COVID-19 pandemic reported high rates of depression, anxiety, and acute stress reactions in healthcare workers in the United States and 14 other countries (Sanghera et al., 2020). These effects are reminiscent of the increased rates of depression, anxiety, and other signs of distress observed in healthcare workers during the SARS and H1N1 epidemics (Busch et al., 2021; Maunder et al., 2006) and highlight the need for large scale interventions that protect healthcare workers from the psychological sequalae of major healthcare crises, which can be long-lasting in some cases (Lee et al., 2018).

Research conducted during these prior epidemics suggest that even brief psychosocial interventions delivered to healthcare workers during such outbreaks can improve outcomes. For example, during the H1N1 epidemic in 2008, an intervention for healthcare workers was developed to focus on education about the anticipated epidemic and potential illnesses, as well as resilience training tools, such as mindfulness and other coping strategies (Maunder et al., 2010). Improvements in self-efficacy, confidence in their training, and reductions in interpersonal problems were reported by healthcare workers after the intervention (Maunder et al., 2010).

Based on this prior work, in March 2020, in anticipation of an imminent surge in COVID-19 cases in the Northeast United States, we developed a brief online course that aimed to introduce employees of a major healthcare system to psychological techniques that might enhance their ability to handle stress. Evidence from prior research conducted in a range of diverse populations suggests that certain skills, such as mindfulness and other emotion regulation and stress management techniques, can increase healthcare workers’ capacity to manage and adapt to stressful circumstances (Kemper et al., 2015). Resilience, defined as the ability to “bounce back” after challenging, highly stressful life events (Rutter, 1985), is thought to be a modifiable capacity and process linked to adaptive outcomes (Choi et al., 2019).

The online course, called Resilience Training (RT) for Healthcare Workers, consists of three 12-20 minute videos that include didactic information, experiential exercises and testimonials from healthcare workers that focus on three evidence-based skills or capacities shown to maintain or increase aspects of emotional resilience: mindfulness (Potes et al., 2018), mentalization (Daubney and Bateman, 2015), and self-compassion (Neff and Germer, 2013). The course materials were adapted from a related, existing program developed for young adults (Burke et al., 2020).

In this study, we first collected baseline information about levels of distress in healthcare workers, as reflected by self-reported anxiety, depression, and worries about the COVID-19 pandemic. In order to make the program rapidly and widely available at the onset of the SARS-CoV-2 outbreak in Boston, we employed a pragmatic non-randomized design, testing whether levels of resilience improved and distress decreased in those healthcare workers who opted to take the course compared to those who did not. In addition, we assessed whether baseline and improvements in resilience correlated with reductions in distress. Lastly, we tested whether exposure to patients with COVID-19 influenced these outcomes, since such exposure has been shown previously to correlate with distress in healthcare workers during the pandemic (Muller et al., 2020).

## Methods

The three-session online RT course was one component of a larger set of wellness offerings for employees within the Massachusetts General Brigham (MGB) healthcare system that included two other online courses, as well as an overall effort to disseminate numerous wellness resources to employees in this healthcare system (see: https://www.massgeneral.org/psychiatry/guide-to-mental-health-resources/general-mental-health-and-coping). Given the immediate need for such materials during the beginning of the COVID-19 pandemic in Boston, Massachusetts, these resources were offered to all employees. The RT course was made available via an online platform (HealthStream™) used to deliver mandatory trainings to employees of the MGB healthcare system. The course was advertised in system-wide emails listing wellness resources for employees. Those who enrolled in the study completed self-report surveys (without compensation) via REDCap at baseline, prior to participation in the online resilience training, then one month and two months later. Neither completion of the surveys nor participation in the course were required; the course was offered to all MGB employees in order to provide unfettered access to any useful support for employees during the COVID-19 crisis. Inclusion criteria included being: 1) a current employee of the MGB Healthcare System, 2) 18 years or older in age. The study protocol was approved by the MGB Institutional Review Board.

### Resilience Training Course

The online RT for healthcare workers course consists of didactic materials delivered in pre-recorded videos by experienced doctoral level clinicians, testimonials of healthcare workers about their experiences during the pandemic and their use of the skills taught in the course, and brief experiential exercises. Session 1 focused on the concept of resilience and mindfulness skills (Potes et al., 2018); session 2 focused on enhancing cognitive flexibility via cognitive behavioral (Beck et al., 1979) and mentalization (Daubney and Bateman, 2015) skills; and session 3 focused on the development of self-compassion (Neff and Germer, 2013). All three sessions emphasized ways to implement these concepts and skills in everyday life, highlighting specific challenges faced by healthcare workers during the COVID-19 pandemic.

### Survey

Participants completed an online 23-item survey three times: at baseline (prior to viewing the course), and one month and two months afterwards. To minimize participant burden, particularly in light of the substantial workload of healthcare workers during the pandemic, the survey was composed of a limited number of items selected from validated questionnaires, with the exception of the Patient Health Questionnaire-4 (PHQ-4; (Löwe et al., 2010), which was included in its entirety (four items, two assessing anxiety and two assessing depression) to capture emotional distress. Other questions assessed a range of outcomes including demographic characteristics, COVID-19 related anxiety, resilience (with two items from the Brief Resilience Scale (Smith et al., 2008)), coping (Park et al., 2021); loneliness (with two items from the UCLA Loneliness Scale (Russell, 1996)); self-compassion (with one item from the Self-Compassion Scale (Neff et al., 2019)), and burn-out (with one item from the Maslach Burnout Inventory (Maslach et al., 1986).

### Statistical Analyses

Our primary analyses focused on two outcomes: 1) resilience (the sum of the ratings of the level of agreement from 1-5 on the following three items: “I am able to bounce back quickly after hard times,” “I am able to come through difficult times with little trouble,” and “I am able to cope with the stress in my life”) and 2) emotional distress (the sum of endorsement ratings on a 1-4 scale on the following four items of the PHQ-4: “feeling nervous, anxious or on edge,” “not being able to stop or control worrying,” “feeling down, depressed, or hopeless,” and “little interest or pleasure in doing things”). Frequencies and Pearson’s correlations were measured first in the baseline sample, then in the longitudinal sample (i.e., those who completed all three assessments) separately. To examine the effect of the RT course on resilience and emotional distress, paired sample *t*-tests were used to compare the baseline and the two longitudinal time points. Repeated measures ANOVAs were then used to assess the two primary outcomes in those who participated in the RT intervention and those who did not. The ANOVA exploring the effects of RT on emotional distress included COVID-19 related anxiety as a covariate since it was significantly associated with emotional distress (*p* < .001).

## Results

The results reported here are based on analyses of the participants who completed the baseline survey (n = 554), those who completed the baseline and second survey (n = 163), and those who completed all three surveys (n = 148). Of those who completed all three surveys, 38 viewed at least one session of the RT course, 28 viewed at least two sessions and 27 viewed all three.

### The Baseline Sample

#### Demographic Characteristics

A total of 554 MGB employees completed the baseline survey (between April 14, 2020 and July 30, 2020). During that time, the number of confirmed COVID-19 cases in Massachusetts increased from 28,163 to 109,400 (Massachusetts Department of Public Health, 2021). Of the baseline participants, 87% reported that they were working in the hospital and 42% reported having at least some contact with COVID-19 patients. Within the three main healthcare worker groups (nurses (26.9%), physicians (13.9%), and other hospital roles (59.2%)), 81% of nurses, 75% of physicians, and 45% of those with other hospital roles were working in-person at their healthcare facility at the time of taking the survey. A total of 68% of nurses, 57% of physicians, and 27% of other workers had had at least one contact with a COVID-19 positive individual. Participant characteristics are reported in Table 1.

**Table 1.**
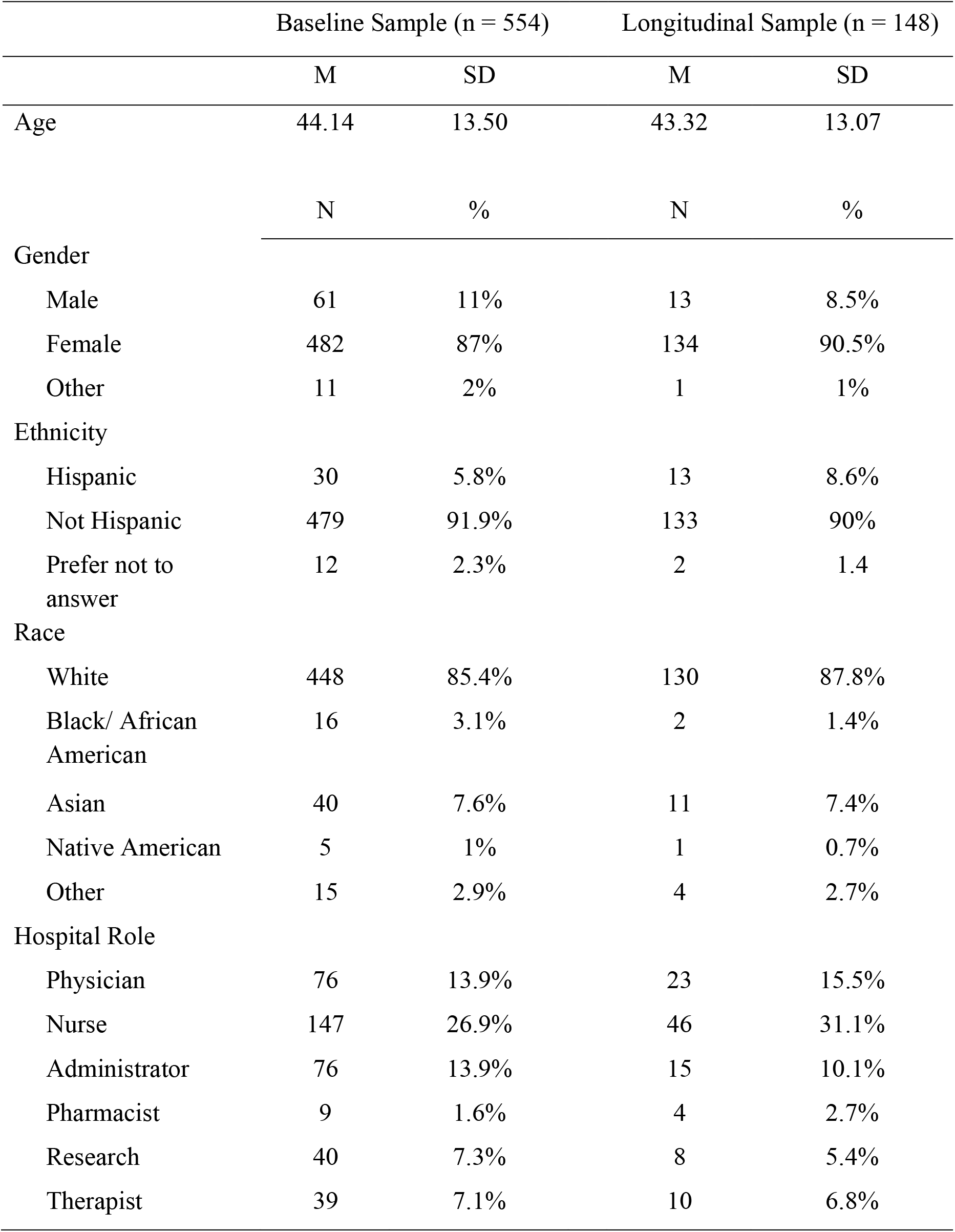
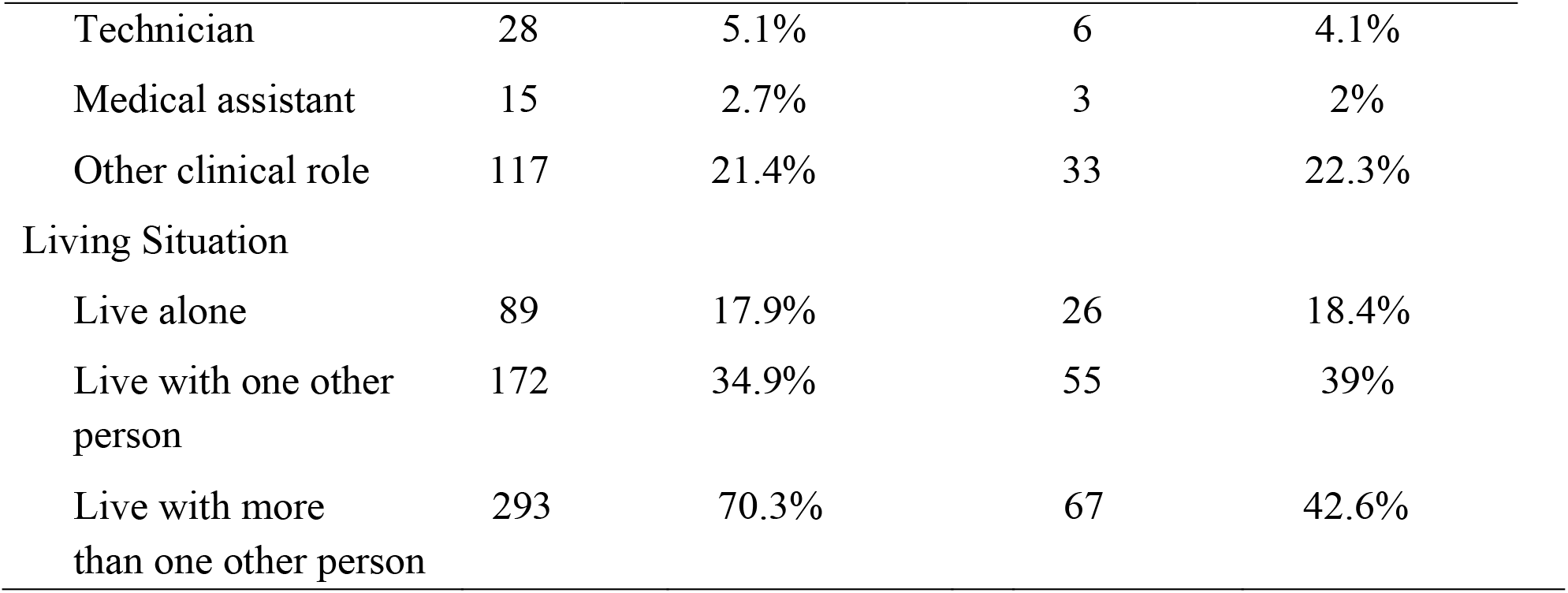
Participant Characteristics

Participant Characteristics
|  | Baseline Sample (n = 554) |  | Longitudinal Sample (n = 148) |  |
| --- | --- | --- | --- | --- |
|  | M | SD | M | SD |
| Age | 44.14 | 13.50 | 43.32 | 13.07 |
|  | N | % | N | % |
| Gender |  |  |  |  |
| Male | 61 | 11% | 13 | 8.5% |
| Female | 482 | 87% | 134 | 90.5% |
| Other | 11 | 2% | 1 | 1% |
| Ethnicity |  |  |  |  |
| Hispanic | 30 | 5.8% | 13 | 8.6% |
| Not Hispanic | 479 | 91.9% | 133 | 90% |
| Prefer not to answer | 12 | 2.3% | 2 | 1.4 |
| Race |  |  |  |  |
| White | 448 | 85.4% | 130 | 87.8% |
| Black/ African American | 16 | 3.1% | 2 | 1.4% |
| Asian | 40 | 7.6% | 11 | 7.4% |
| Native American | 5 | 1% | 1 | 0.7% |
| Other | 15 | 2.9% | 4 | 2.7% |
| Hospital Role |  |  |  |  |
| Physician | 76 | 13.9% | 23 | 15.5% |
| Nurse | 147 | 26.9% | 46 | 31.1% |
| Administrator | 76 | 13.9% | 15 | 10.1% |
| Pharmacist | 9 | 1.6% | 4 | 2.7% |
| Research | 40 | 7.3% | 8 | 5.4% |
| Therapist | 39 | 7.1% | 10 | 6.8% |
| Technician | 28 | 5.1% | 6 | 4.1% |
| Medical assistant | 15 | 2.7% | 3 | 2% |
| Other clinical role | 117 | 21.4% | 33 | 22.3% |
| Living Situation |  |  |  |  |
| Live alone | 89 | 17.9% | 26 | 18.4% |
| Live with one other person | 172 | 34.9% | 55 | 39% |
| Live with more than one other person | 293 | 70.3% | 67 | 42.6% |

#### Associations

Across all participants, moderate to high levels of baseline emotional distress (M = 8.15, SD = 2.92 on a scale of 1-12, with scores of 6-8 on the PHQ-4 representing “moderate” levels of anxiety and depression (Löwe et al., 2010)) were found. Loneliness scores revealed that participants endorsed feeling lonely “several days” per week on average (M = 1.7, SD = .77). Exposure to patients with COVID-19 did not correlate with baseline emotional distress or loneliness (*r* = .046 and .058, respectively). However, in the full sample, greater exposure to COVID-19 patients was associated with a greater likelihood of feeling that one has positively impacted others (*r* = .171, *p* < .001). A feeling of positively impacting others was also significantly related to lower emotional distress (*r* = -.285, *p* < .001). Lastly, significant correlations were found between emotional distress, loneliness, COVID-19-related anxiety, and resilience (see Table 2).

**Table 2.** Baseline Correlations (N = 554, Pearson’s R Values)

|  | Emotional Distress | Loneliness | COVID-19<br>Related Anxiety |
| --- | --- | --- | --- |
| Emotional Distress |  |  |  |
| Loneliness | .539** |  |  |
| COVID-19 Related Anxiety | .275** | .264** |  |
| COVID-19 Exposure | .046 | .058 | .100* |
| Resilience | -.535** | -.304** | -.138 |
| Self-Compassion | -.334** | -.202** | -.162** |
| Positively Impacting Others | -.284** | -.168** | -.058 |

Note: \* $p < .05$ ; \*\* $p < .001$ .

### The Longitudinal Sample

#### Demographic Characteristics

A total of 148 participants completed all three assessments. The mean dates-of-completion for the second and third time point were June 20, 2020 and July 31, 2020, respectively. A total of 86% of this sample was working in-person during this time and 44.6% had contact with a COVID-19 patient. The demographic characteristics of this sample were similar to those of the baseline sample (see Table 1).

#### Associations of Baseline Resilience with Longitudinal Outcomes

Greater levels of baseline resilience were associated with lower levels of emotional distress (rated on the PHQ-4) at both the second (*r* = -.407, *p* < .001) and third (*r* = -.445, *p* < .001) timepoints. Similar relationships with baseline resilience were observed for loneliness at the second (*r* = -.384, *p* < .001) and third (*r* = -.406, *p* < .001) timepoints. Greater exposure to COVID-19 patients at baseline was not related to emotional distress at subsequent time points (all *p*s > .05).

#### Effects of the RT Course within Subjects

A total of 231 participants (of 554 (41.7%)) viewed at least a portion of the RT course, with 115 (of 554 (20.7%)) viewing all three sessions. Among those who took the RT course and completed all of the assessments (n = 38), there was no significant change in resilience levels from baseline to one month following the course (*t* = 1.46, *p* = .153) but subsequently, resilience levels significantly increased from baseline to two months following the course (*t* = 2.88, *p* = .010). In addition, significant decreases in emotional distress were observed at one month (*t* = 3.09, *p* = .004) and two months following the course (*t* = 2.97, *p* = .009).

#### Comparisons between those who took the RT Course and those who did not

Among the 148 participants who completed all three assessments, there were no significant differences in demographic variables, or in baseline resilience and emotional distress, between those who took the RT course (n = 38) compared to those who did not (n = 110) (all *p*s > .10). A repeated measure ANOVA testing for effects of the RT course on resilience revealed a significant group by time interaction (*F*(2, 122) = 3.56, *p* = .031); those who took the RT course showed sustained improvements in resilience over the two month follow-up period compared to those who did not (Figure 1). Similarly, a repeated measures ANOVA testing for effects of the RT course on emotional distress also revealed a significant group by time interaction (*F*(2, 116) = 3.15, *p* = .047), due to reductions in emotional distress in the RT participants compared to those who did not take the course (Figure 2).

**Figure 1.**
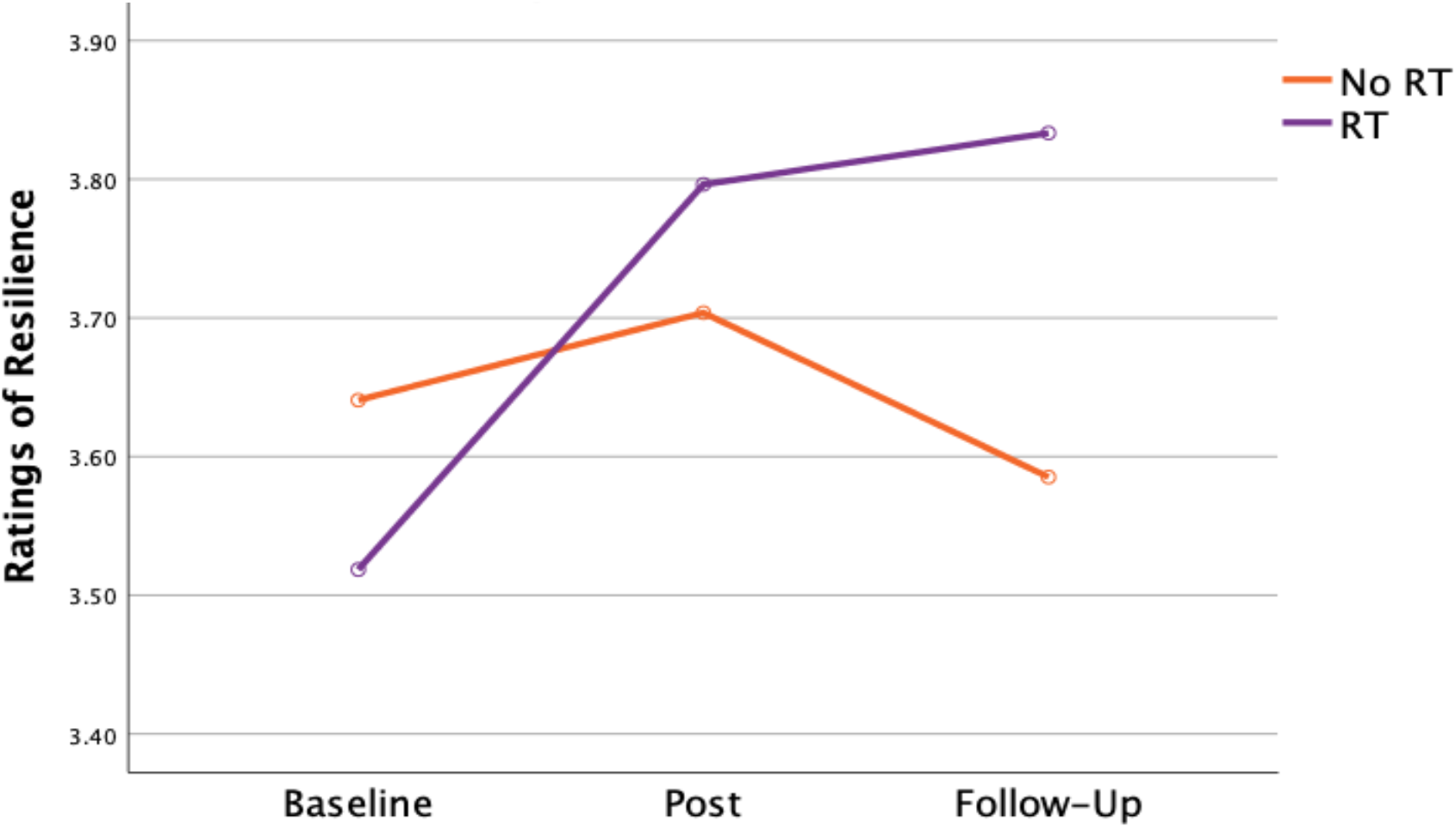
Resilience Levels of Those Who Participated in the Resilience Training (RT) Course (n = 38) and Those Who Did Not (n = 110). Self-reported resilence, as measured using a 4-item composite scale, improved in the MGB healthcare workers who chose to take the Resilience Training (RT) for Healthcare Workers 3-session online course over the two month follow-up period, compared to those who did not, as reflected by a significant group by time interaction = *F*(2, 122) = 3.562, *p* = .031.

**Figure 2.**
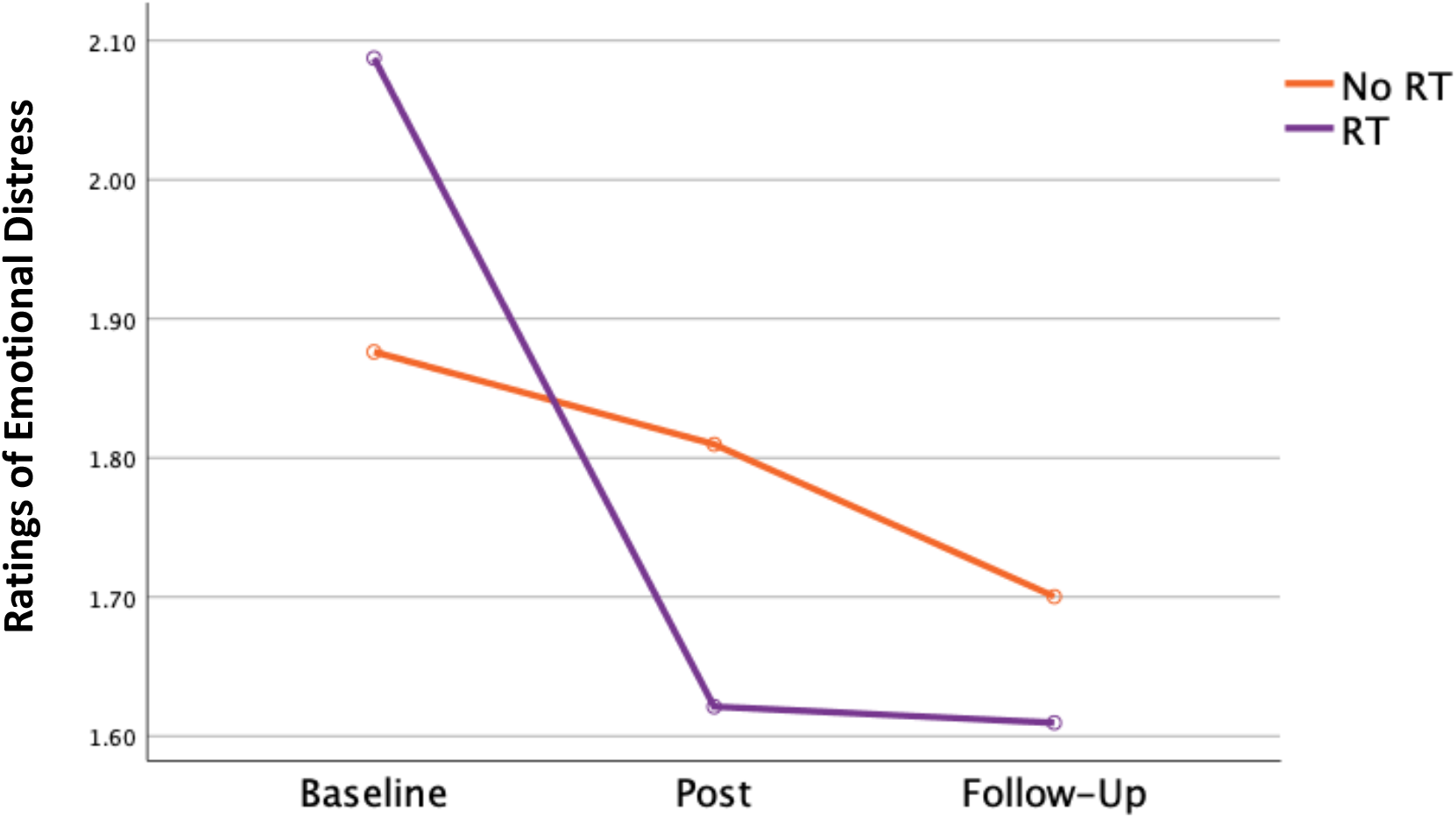
Emotional Distress Levels in Those Who Participated in the Resilience Training (RT) Course (n =38) and Those Who Did Not (n = 110). Self-reported levels of anxiety and depression, as measured using the 4-item PHQ-4 scale, decreased in the MGB healthcare workers who chose to take the Resilience Training (RT) for Healthcare Workers 3-session online course over the two month follow-up period, compared to those who did not, as reflected by a significant group by time interaction *F*(2,116) = 3.145, *p* = .047, with COVID-19 worry included as a covariate.

Consistent with these results, within the group that took the RT course, improvements in resilience correlated with decreases in emotional distress both at one month (*r* = -.560 *p* < .001) and two months (*r* = -.666, *p* = .003) following the course.

## Discussion

This study replicates findings of other studies of the mental health of healthcare employees during the COVID-19 pandemic (Lai et al., 2020) and prior pandemics (Busch et al., 2021; Maunder et al., 2006), which have shown that this vulnerable population experiences moderate to high levels of distress and a sense of isolation during such crises. In addition, in the intervention portion of the study, which by necessity used a pragmatic, nonrandomized design, improvements in resilience and decreases in emotional distress were observed in healthcare employees who chose to participate in the brief resilience training course. These initial findings, while they must be interpreted with caution given the small size of the sample and non-randomized design, further support the possibility that a brief, online intervention can be beneficial to healthcare workers even in the midst of a crisis (Maunder et al., 2010), potentially providing a much-needed boost to mental health during a time of extraordinary stress. Moreover, these results are consistent with prior evidence that healthcare workers are willing to engage in online, wellness-focused courses (Blake et al., 2020), and such courses can be effective and helpful for this population (Maunder et al., 2010).

Of note, surprisingly (Muller et al., 2020) exposure to COVID-19 patients was not significantly correlated with emotional distress, suggesting that the majority of the participants may have felt the psychological impact of the pandemic regardless of whether or not they had had direct exposure to a patient infected with the virus. During the first surge of the pandemic when testing was infrequent (when much of these data were collected), healthcare workers may have been highly aware of the increased risk of infection for themselves and their family members that was associated with their jobs (Adams & Walls, 2020; Black et al., 2020). Thus, these healthcare employees may have been generally impacted by knowledge of this risk, independent of their degree of COVID-19 exposure.

However, greater exposure to COVID-19 patients was associated with a greater likelihood of feeling that one was positively impacting others, suggesting that being involved in the care of such patients may have been psychologically protective to some extent, potentially conferring a greater sense of purpose and meaning during the crisis. This is consistent with prior evidence for links between having a sense of purpose or meaning in one’s life and greater resilience (Ostafin and Proulx, 2020).

All three of the skills taught in the RT intervention, mindfulness, mentalization, and self-compassion, have been shown to decrease various forms of emotional distress, such as symptoms of anxiety and depression (Frostadottir and Dorjee, 2019; Hayden et al., 2018; Hofmann and Gómez, 2017). These skills were focused on in the course in order to foster key emotion regulation capacities, thereby allowing healthcare workers to engage in a type of active resilience building (Kalisch et al., 2015). They were also selected because of the evidence for their positive effects on social functioning and connection (Burke et al., 2020; Lindsay et al., 2019). Given the increased social isolation experienced by many during the COVID-19 pandemic, including healthcare workers who have been often required to work long hours in masks and other protective equipment while maintaining “social distance” from others, augmenting capacities that may lead to more meaningful connections with others may help to preserve well-being during this unusual period and its aftermath, as well as during other times of adversity.

In light of the need for such interventions for healthcare workers both during and following the COVID-19 pandemic, programs like this merit further study while being simultaneously rapidly implemented. In designing follow-up work, several limitations of this study should be considered. The non-randomized design of this study, although necessary given the high level of need of the sample, limits the inferences that can be made from these findings. Also, enrollment was likely affected by the fact that these healthcare employees were being flooded daily with lengthy emails regarding COVID-19-related information and supports, some of which included information about this study. This “information overload”, and the other multiple new demands associated with the crisis, may have also interfered with the capacity of participants to both view the videos and complete all three assessments.

In summary, this study provides preliminary evidence for the efficacy of a brief online intervention for improving resilience and decreasing emotional distress in healthcare workers during the COVID-19 pandemic. Because some healthcare workers will likely need ongoing, additional support after the pandemic has subsided, resilience-enhancing interventions that are tailored to meet the needs of this population at subsequent points in time should also be developed and implemented.

## Data Availability

The data that support the findings of this study are available from the corresponding author, Dr. DeTore, upon reasonable request.

## Acknowledgements

We would like to thank a number of critical contributors to the development of the online intervention and data collection (including healthcare workers and experts who provided testimonials or advice within the course, members of the film crew, and research staff, listed alphabetically): Amber Leonard Alibrio, Archana Basu, Wisteria Deng, Brad Dickerson, Darcie Edwards, Rich Fomo, Nevita George, Chris Germer, Chad Gobert, Diana Johnson, Rachel Kakos, Megan Koster, Seth Margolis, Mia Mazzaferro, Roberto Mighty, Julie Newbold, Olivia Okereke, Steven Pugliese, Phoebe Ramler, Christopher Richard, Matthew Robinson, Nancy Rotter, Lauren Schiffner, Lauren Washington, Lynn Weissman, Suzanne Willard-Kiess, Sarah Zapetis, and Janet Zedler.

